# CaPri: Bayesian Mendelian randomization for joint estimation of causal and pleiotropic effects

**DOI:** 10.64898/2026.10.13.26365454

**Authors:** Ou Ren, Wenmin Zhang, Tianyuan Lu

## Abstract

**Motivation:** Horizontal pleiotropy remains a major challenge for Mendelian randomization (MR), particularly as increasingly large genome-wide association studies identify many genetic instruments for complex traits. Existing pleiotropy-robust methods rely on assumptions regarding the proportion or distribution of valid instruments that may be violated when pleiotropy is widespread.

**Results:** We developed CaPri, a Bayesian MR framework that jointly estimates causal and pleiotropic effects. CaPri allows a potentially arbitrary proportion of instruments to contribute to a shared directional pleiotropic component and estimates the posterior probability and magnitude of the causal effect, instrument-specific posterior inclusion probabilities for pleiotropy, and the shared pleiotropic effect. Across simulations, CaPri maintained appropriate false positive control and accurate causal effect estimation regardless of the proportion of pleiotropic instruments, while accurately identifying pleiotropic instruments and estimating their shared effect. Performance remained robust across varying causal and pleiotropic effect sizes and exposure-outcome confounding, although sufficiently large outcome GWAS sample sizes were important for stable inference and limited prior dependence. In real data analyses, CaPri reported low posterior probabilities of causality across negative controls and strong support for established positive control relationships. Application to alcohol consumption and coronary artery disease further demonstrated its ability to identify potentially meaningful pleiotropic instruments. CaPri provides a complementary framework for strengthening causal inference and triangulation across MR methods with different identifying assumptions.

**Availability and implementation:** CaPri is available at https://github.com/OuRen0104/CaPri.

## Introduction

Mendelian randomization (MR) has become widely used to evaluate the potential causal effects of modifiable exposures on health outcomes using genetic variants as instrumental variables^1^. By leveraging the random allocation of genetic variants at conception, MR can be less susceptible to conventional confounding and reverse causation than observational analyses^2^. Valid causal inference using MR relies on three core instrumental variable assumptions^3,4^: genetic instruments must be associated with the exposure of interest; the instrument-outcome association must not be confounded; and the instruments must affect the outcome only through the exposure. The first assumption, commonly referred to as the relevance assumption, can generally be addressed by selecting genetic variants that are strongly associated with the exposure. The second assumption, often referred to as the independence assumption, is supported by the random allocation of genetic variants at conception. Nevertheless, population stratification, selection bias, assortative mating, dynastic effects, and other sources of residual confounding can induce associations between genetic instruments and the outcome that do not reflect a causal pathway through the exposure^5–8^. These potential biases can be substantially reduced through careful genome-wide association study (GWAS) design and analysis. In contrast, the third assumption, known as the exclusion restriction assumption, remains particularly difficult to assess because horizontal pleiotropic effects of genetic variants cannot be directly ruled out.

This challenge has become increasingly important as MR is applied to highly polygenic exposures. Large-scale GWAS now identify tens to hundreds of independent genetic instruments for many complex traits, substantially increasing statistical power but also increasing the opportunity for individual variants to influence the outcome through pathways independent of the exposure^9–12^. Conventional inverse-variance weighted MR assumes that all included genetic variants are valid instruments, and violations of this assumption due to horizontal pleiotropy can lead to biased causal estimates^13^. Growing empirical evidence suggests that pleiotropy is widespread across complex traits^14^, making the exclusion restriction assumption increasingly difficult to justify as larger sets of genetic instruments are used.

Several methods have therefore been developed to provide robust causal estimates in the presence of invalid or pleiotropic instruments, but each relies on additional assumptions about the underlying structure of pleiotropy. MR Egger regression allows all instruments to have direct effects on the outcome but requires the InSIDE (Instrument Strength Independent of Direct Effect) assumption, under which the magnitude of an instrument’s association with the exposure is independent of its direct effect on the outcome^15^. Median-based estimators remain consistent when at least half of the information contributing to the analysis comes from valid instruments^16,17^, whereas mode-based estimators require the largest cluster of instruments with similar causal estimates to consist of valid instruments^18^. MR PRESSO identifies and removes variants with outlying pleiotropic effects and performs best when horizontal pleiotropy is restricted to a minority of instruments^19^. These approaches provide important strategies for addressing horizontal pleiotropy, but their identifying assumptions may not hold for highly polygenic traits, for which pleiotropy may involve a substantial proportion of genetic instruments.

In this study, we introduce CaPri, a Bayesian framework for MR that jointly models the causal effect of an exposure and directional pleiotropic effects of its genetic instruments. CaPri uses a probabilistic graphical model in which each instrument is assigned a posterior probability of contributing to a shared pleiotropic effect, allowing any proportion of the genetic instruments to be pleiotropic under the specified model. We developed an efficient variational inference algorithm^20–22^ to jointly estimate the posterior probability and magnitude of the causal effect, as well as instrument-specific posterior probabilities of pleiotropy and the shared pleiotropic effect. We evaluated CaPri through extensive simulations across varying proportions of pleiotropic instruments, causal effect sizes, pleiotropic effect sizes, and strengths of exposure-outcome confounding, and examined its robustness to alternative prior specifications and outcome GWAS sample sizes. We further evaluated its practical utility in real data analyses using negative control and established positive control exposure-outcome pairs, followed by an application to the relationship between alcohol consumption and coronary artery disease.

## Materials and methods

### Model overview

The probabilistic graphical model underlying CaPri is illustrated in **Figure 1**. Consider *K* independent genetic instruments. For the *k*-th instrument, let 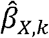 and 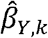 denote its estimated associations with the exposure and outcome, respectively. CaPri introduces a binary indicator *δ* for whether the exposure has a causal effect on the outcome, and denotes the causal effect, with γ = 0 when δ = 0. For each instrument, *c*_*kY*_ is a binary indicator of whether the instrument has a direct pleiotropic effect on the outcome, and β_*Y*_ denotes the shared pleiotropic effect among instruments with *c*_*kY*_ =1.

**Figure 1.**
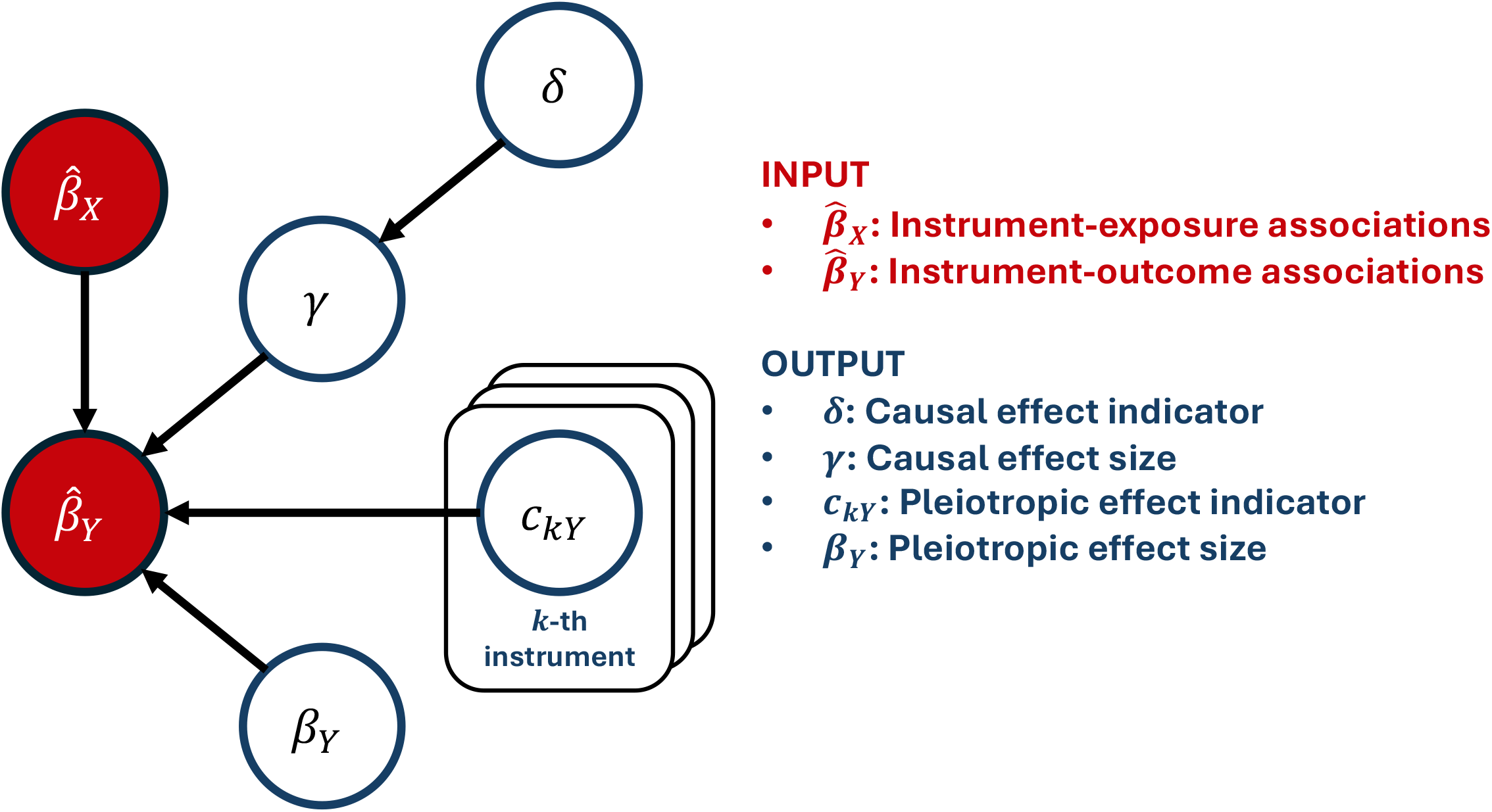
Probabilistic graphical model underlying CaPri. 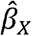 and 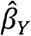 denote the estimated instrument-exposure and instrument-outcome associations from genome-wide association studies, respectively. δ indicates whether the exposure has a causal effect on the outcome, *γ* and denotes the causal effect. *c*_*kY*_ indicates whether the *k*-th instrument contributes to the pleiotropic component, and *β*_*Y*_ denotes the shared pleiotropic effect. CaPri models the instrument-outcome association as the combination of a causal component and a shared pleiotropic component contributed by instruments with *c*_*kY*_ = 1.

CaPri models the instrument-outcome association as the combination of a causal component and a pleiotropic component:

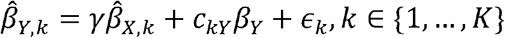

where 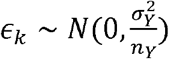 with 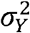 representing the variance in the outcome not explained by the instruments and *n*_*Y*_ representing the outcome GWAS sample size. The instrument-exposure associations 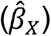 are treated as fixed.

For the causal component, we assign

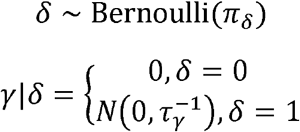

For the pleiotropic component,

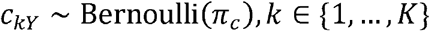

and the shared pleiotropic effect is assigned the prior kernel

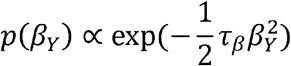

When τ_β_> 0, this corresponds to 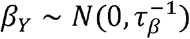, whereas τ_β_= 0 corresponds to a flat, non-informative prior on 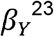.

The primary quantities of interest are the posterior probability that the exposure has a causal effect on the outcome, *P*(o 1|data), denoted PP.causal; the posterior mean causal effect; the posterior inclusion probability of each instrument in the pleiotropic component, *P*(c_*kY*_ =1|data), denoted PIP; and the posterior mean shared pleiotropic effect. CaPri uses coordinate-ascent variational inference to jointly approximate these posterior distributions, enabling efficient inference when a large number of genetic instruments are included. Full derivations and the variational inference algorithm are provided in the **Supplementary Notes**.

We provide default prior settings that are intended to impose limited prior constraints. We set *π*_*δ*_=0.01, corresponding to a weak prior probability that the exposure has a causal effect on the outcome, and *τ*_*γ*_ *=* 1, providing modest regularization of the causal effect size. We set *π*_*c*_=0.5, assigning each instrument an equal prior probability of being pleiotropic or non-pleiotropic. For the shared pleiotropic effect, we set *τ*_*β*_ *=* 0. Finally, we set 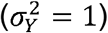 throughout this study because the outcomes were standardized and the genetic instruments explained a relatively small proportion of the outcome variance, such that the residual variance was expected to be close to one. In applications with an unstandardized outcome, 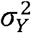 can be specified according to the residual variance of the outcome. The sensitivity of CaPri to these prior specifications was evaluated in additional analyses described below.

### Simulation study design

We performed simulation studies to evaluate the performance of CaPri under varying proportions of pleiotropic instruments, causal effect sizes, pleiotropic effect sizes, strengths of exposure-outcome confounding, and outcome GWAS sample sizes. We simulated two independent samples representing the exposure and outcome GWAS. We simulated *K* = 20 genetic instruments, with exposure and outcome GWAS sample sizes *N*_*x*_*=* 50000 and *N*_*Y*_*=* 50000, respectively; the proportion of exposure variance attributable to the genetic component 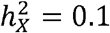 causal component parameter 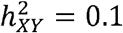 direct pleiotropic component parameter 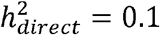; and confounding parameters 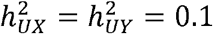. We varied the number of pleiotropic instruments as *K*_*pleiotropiC*_ *∈* {0,5,10,15,20}. In additional simulations, we varied the magnitude of the causal component, 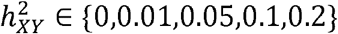; the magnitude of the pleiotropic component, 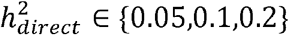 the strength of exposure-outcome confounding, with 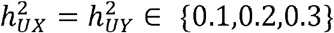 and the outcome GWAS sample size, *nY ∈ {*2000,5000,10000}. All simulation settings are summarized in **Supplementary Table 1**. Each simulation setting was independently replicated 100 times.

For each genetic instrument, the minor allele frequency was sampled from a uniform distribution,

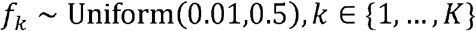

and genotype dosages for the *i*-th individual were independently generated in the exposure and outcome samples as

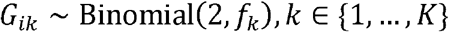

We next generated |α_*X,k*_| ~ |*N*(0,1)| +3. The same minor allele frequencies and underlying instrument-exposure effects were used in the two samples. We then constructed a standardized genetic component with zero mean and unit variance for the exposure,

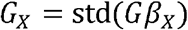

Within each sample, we independently generated a latent variable *U*~ *N*(0,1) to induce confounding of the exposure-outcome relationship. The standardized exposure was generated as

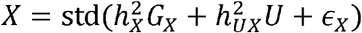

where 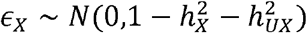

Instrument-exposure associations, 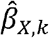, and their standard errors were estimated in the exposure GWAS sample using separate marginal linear regressions of *X* on each genetic instrument.

In the outcome GWAS sample, *K*_*pleiotropiC*_ instruments were randomly selected to have direct pleiotropic effects. The pleiotropic genetic component was defined as

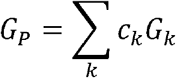

When at least one pleiotropic instrument was present, the standardized outcome was generated as

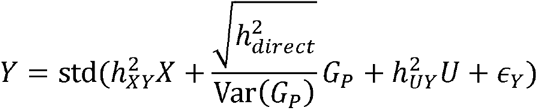

where 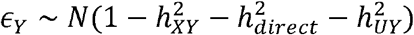.

When no pleiotropic instruments were present, the direct pleiotropic component was omitted and 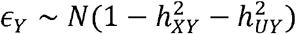.

Instrument-outcome associations, 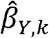, and their standard errors were estimated using separate marginal linear regressions of *Y* on each genetic instrument.

Because standardization changes the numerical scale of the simulated effects, we estimated ground-truth causal and pleiotropic effect sizes from the realized individual-level data.

When pleiotropy was present, we fitted

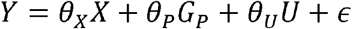

and used the estimated coefficients 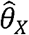 and 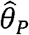 as the ground-truth causal and pleiotropic effects, respectively. When 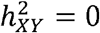, the ground-truth causal effect was set to zero.

When no pleiotropic instruments were present, we fitted

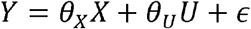

and used the estimated coefficient 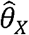 as the ground-truth causal effect, while the ground-truth pleiotropic effect was set to zero.

### Performance evaluation in simulation study

We compared CaPri with conventional and pleiotropy-robust MR methods, including inverse-variance weighted (IVW) MR using the default random-effects model, fixed-effects IVW, simple median, weighted median, penalized weighted median, simple mode, weighted mode, MR Egger regression, and MR PRESSO. All MR methods, except MR PRESSO, were implemented using the TwoSampleMR R package (version 0.7.0), while MR PRESSO was implemented using the MRPRESSO R package (version 1.0).

We first evaluated causal effect detection and estimation. When a causal effect was present, we calculated the proportion of simulation replicates in which CaPri reported PP.causal > 0.95, compared with the proportion with P < 0.05 for other MR methods. We additionally evaluated more stringent thresholds of PP.causal > 0.99 for CaPri and P < 0.01 for other methods. When no causal effect was present, false positive rates were defined as the proportion of simulation replicates with PP.causal > 0.05 for CaPri and P < 0.05 for other methods. This permissive threshold for CaPri provides a stringent assessment of false positive control. Causal effect estimation error was calculated as the absolute difference between the estimated and ground-truth causal effects.

We next evaluated the identification and estimation of pleiotropic effects. For CaPri, an instrument was classified as pleiotropic when its PIP exceeded 0.95, with PIP > 0.99 evaluated as a more stringent threshold. We calculated sensitivity, specificity, and precision for identifying the known pleiotropic instruments used in data generation; when no pleiotropic instruments were present, only specificity was evaluated. Pleiotropic effect estimation error was calculated as the absolute difference between the estimated and ground-truth shared pleiotropic effects. For comparison, we evaluated the ability of the MR Egger intercept test to detect directional pleiotropy using P < 0.05 and P < 0.01, and calculated its pleiotropic effect estimation error as the absolute difference between the MR Egger intercept and the ground-truth shared pleiotropic effect.

### Assessment of sensitivity to prior specification

We assessed the sensitivity of CaPri to alternative prior specifications using the same simulated datasets generated under the primary simulation settings. We varied one prior parameter at a time while holding the remaining parameters at their default values.

For the prior probability of a causal effect, we varied *πδ* ∈ {0.0001,0.0005,0.001,0.005,0.01,0.05,0.1,0.5}. For the prior precision of the causal effect, we varied τ_*γ*_ ∈ {0.01,0.05,0.1,0.5,1,5,10}. For the prior precision of the shared pleiotropic effect, we varied τ_*β*_ ∈{0,0.01,0.05,0.1,0.5,1,5,10}

For each prior specification, we refitted CaPri to the same simulation replicates and evaluated changes in PP.causal, the estimated causal effect, and the estimated pleiotropic effect.

### Negative control analyses

We further evaluated CaPri using exposure-outcome pairs selected to represent negative controls, established positive controls, and an exploratory setting. We first performed negative control analyses to evaluate CaPri in real GWAS data, where complex genetic architectures and horizontal pleiotropy may not be fully represented by simulations. We used self-reported ease of skin tanning as the negative control outcome and evaluated ten exposures representing diverse biological and behavioral domains: asthma^24^, Alzheimer’s disease^25^, neuroticism^26^, schizophrenia^27^, body mass index^28^, waist-to-hip ratio^29^, type 2 diabetes^30^, cardiovascular disease^31^, lung cancer^32^, and years of schooling^33^. These exposures were selected because none has a plausible direct biological or mechanistic causal effect on an individual’s inherent tendency to tan^34^. Summary statistics for ease of skin tanning were obtained from a GWAS of up to 378,364 European-ancestry participants in the UK Biobank^14,35^. Ease of tanning was self-reported on a four-level scale ranging from “get very tanned” to “never tan, only burn.”

### Positive control analyses

We next evaluated three exposure-outcome pairs with well-established causal or biological relationships: low-density lipoprotein (LDL) cholesterol and coronary artery disease (CAD), serum urate and gout, and insulin-like growth factor 1 (IGF-1) and height. LDL cholesterol is an established causal determinant of CAD, supported by well-characterized molecular mechanisms, extensive observational evidence, and randomized controlled trials of LDL-lowering medications^36–39^. Summary statistics for LDL cholesterol were obtained from a meta-analysis of up to 188,577 European-ancestry participants from 60 studies in the GLGC Consortium^40^. CAD summary statistics were obtained from a large meta-analysis of European-ancestry participants from the CARDIoGRAMplusC4D Consortium and eight additional studies, including up to 181,522 cases and 984,168 controls^41^.

Elevated serum urate has a well-established biological role in gout through the formation and deposition of monosodium urate crystals^42^. Serum urate summary statistics were obtained from a GWAS of up to 363,228 unrelated UK Biobank participants^11^. Gout summary statistics were obtained from a meta-analysis of 14 studies of European-ancestry participants, including up to 2,115 cases and 67,259 controls^43^. IGF-1 is a major mediator of the growth hormone axis and has a well-established biological role in linear growth^44,45^. Summary statistics for serum IGF-1 were obtained from a GWAS of up to 363,228 unrelated UK Biobank participants^11^. Height summary statistics were obtained from a meta-analysis of up to 253,288 European-ancestry participants from 79 studies in the GIANT Consortium^46^.

### Exploratory analysis of alcohol consumption and coronary artery disease

Finally, we evaluated alcohol consumption and CAD as an exploratory analysis. The causal relationship between alcohol consumption and CAD remains uncertain, particularly at low to moderate levels of consumption. Although observational studies have historically suggested potentially protective associations of moderate alcohol consumption with cardiometabolic disease^47^, these findings may be influenced by confounding and reverse causation^48–50^, and long-term randomized trials of alcohol consumption are difficult to conduct. This exposure-outcome pair therefore provides a setting in which different MR approaches may produce differing conclusions and in which the identification of potential pleiotropic instruments may be particularly informative.

Summary statistics for alcohol consumption were obtained from a GWAS meta-analysis of up to 537,349 European-ancestry participants from 33 studies in the GSCAN Consortium^51^. Alcohol consumption was quantified as the average number of drinks per week and log-transformed before association testing.

## Results

### CaPri accurately estimates causal effects in the presence of widespread pleiotropy

We first evaluated false positive control under the simulation setting with no causal effect 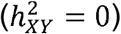. PP.causal for CaPri remained below 0.01 in all simulation replicates, regardless of whether 0, 5, 10, 15, or all 20 instruments had direct pleiotropic effects (**Figure 2A; Supplementary Table 1**).

**Figure 2.**
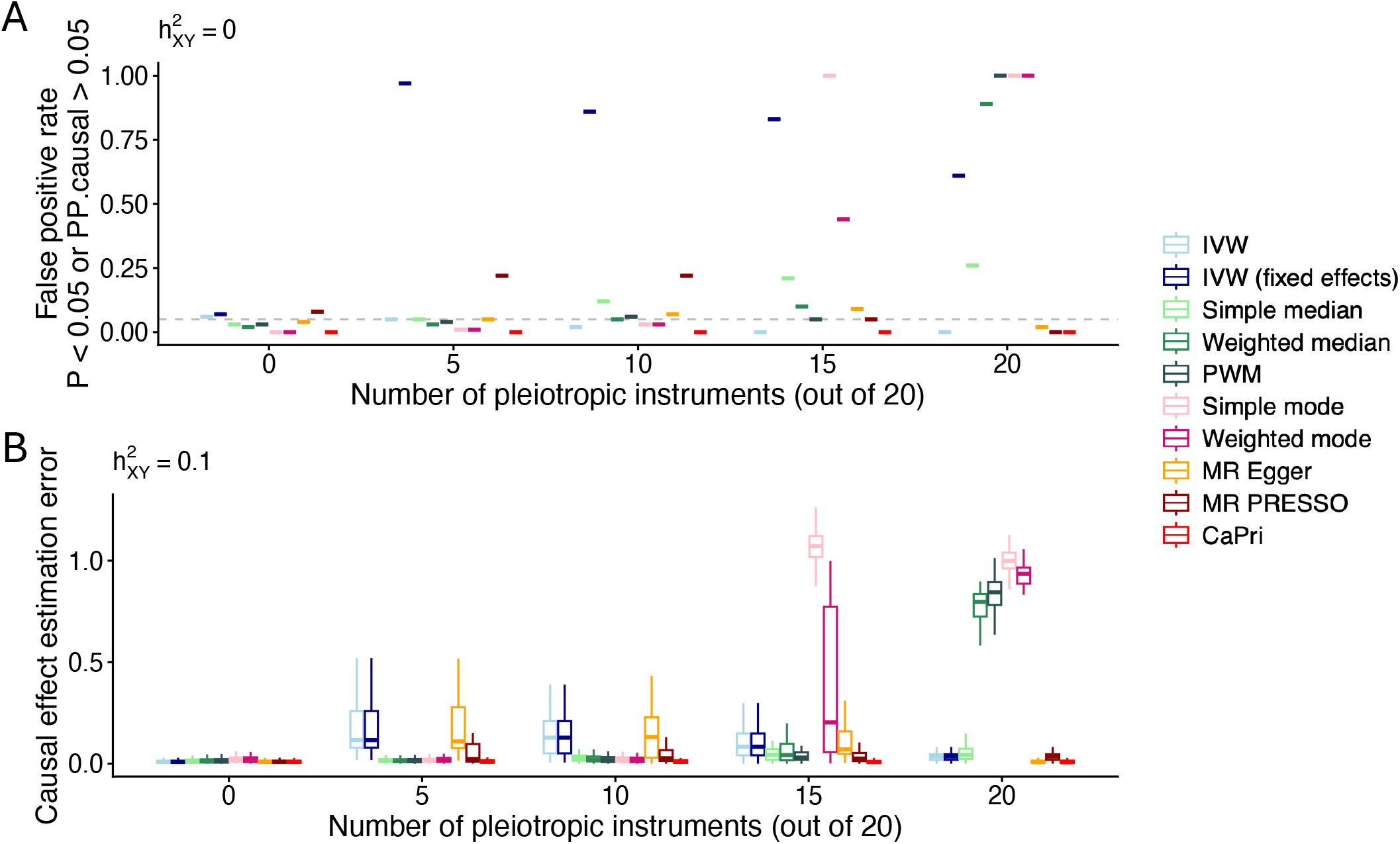
Comparison of CaPri and other Mendelian randomization methods for causal effect inference. Results were obtained from the primary simulation setting with 100 replicates per scenario. Results are shown across simulations with 0, 5, 10, 15, or 20 pleiotropic instruments among 20 genetic instruments. (A) False positive rates under simulations with no causal effect. For CaPri, a false positive result was defined as PP.causal >0.05; for other MR methods, a false positive result was defined as P<0.05. (B) Causal effect estimation error, calculated as the absolute difference between the estimated and ground-truth causal effects. Boxplots show the median (horizontal line), interquartile range (box), and values within 1.5 times the interquartile range from the lower and upper quartiles (whiskers).

In contrast, several conventional MR methods showed increasing false positive rates as pleiotropy became widespread (**Figure 2A; Supplementary Table 1**). This was particularly evident for median- and mode-based methods when more than half of the instruments were pleiotropic. With 15 pleiotropic instruments, the false positive rates were 21% for the simple median, 10% for the weighted median, 100% for the simple mode, and 44% for the weighted mode. When all 20 instruments were pleiotropic, the corresponding rates increased to 26%, 89%, 100%, and 100%, respectively, with the penalized weighted median also reaching 100%. MR PRESSO showed elevated false positive rates in some settings, reaching 22% when 5 or 10 of the 20 instruments were pleiotropic. These findings are consistent with violations of the identifying assumptions of these methods.

We next evaluated causal effect estimation when a true causal effect was present. In the absence of pleiotropy, all methods estimated the causal effect relatively accurately (**Figure 2B; Supplementary Table 1**). As the number of pleiotropic instruments increased, CaPri continued to accurately estimate the causal effect. Under the primary simulation setting 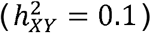, PP.causal exceeded 0.99 in all replicates across all proportions of pleiotropic instruments, and the causal effect estimation error remained low.

In comparison, the accuracy of median- and mode-based estimators deteriorated substantially when pleiotropic instruments constituted the majority (**Figure 2B; Supplementary Table 1**). MR Egger showed a distinct pattern. When all 20 instruments were pleiotropic, its causal effect estimation error was comparable to that of CaPri, as the simulated pleiotropic architecture closely corresponded to the MR Egger model, with all instruments sharing a common directional pleiotropic effect. However, when the shared pleiotropic effect was present in only a subset of instruments, MR Egger was less accurate.

The performance of CaPri was consistent across a broad range of simulation settings (**Supplementary Table 1**). Similar causal effect detection and estimation were observed when varying the magnitude of the causal effect (**Supplementary Figure 1**), the magnitude of the pleiotropic effect (**Supplementary Figure 2**), and the strength of exposure-outcome confounding (**Supplementary Figure 3**). Even with a relatively small causal effect 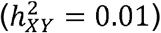, CaPri reported PP.causal > 0.95 in 97%-99% of simulation replicates across different proportions of pleiotropic instruments, increasing to 100% for larger causal effects.

### CaPri accurately identifies and estimates pleiotropic effects

We next evaluated whether CaPri could identify individual instruments contributing to the pleiotropic component. When no pleiotropic instruments were present, instrument-specific PIP were concentrated around 0.5 (**Figure 3A**), corresponding to the prior probability of pleiotropy (*π*_*c*_ = 0.5). Consequently, very few instruments exceeded the prespecified PIP thresholds, with specificity exceeding 0.99 using PIP > 0.99 under the primary simulation setting.

**Figure 3.**
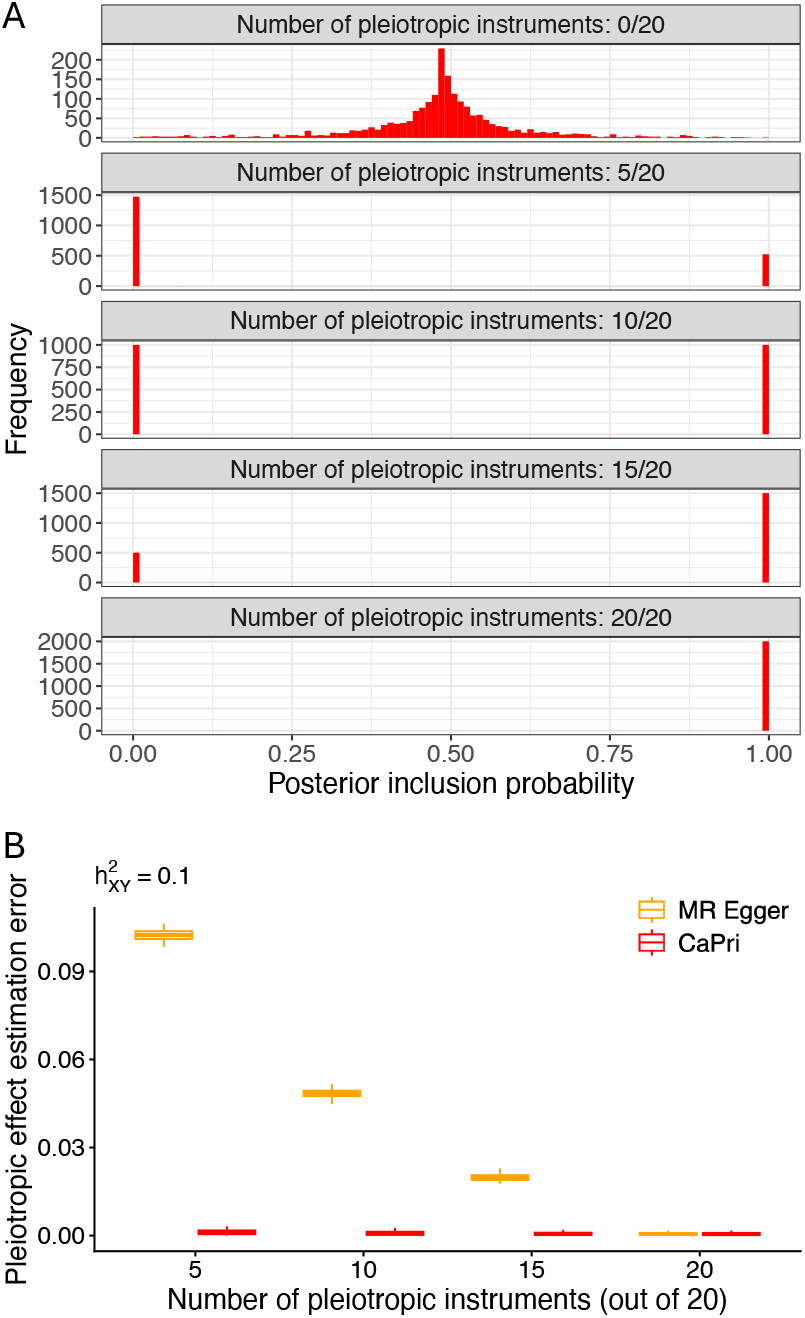
Estimation of pleiotropic effects by CaPri. Results were obtained from the primary simulation setting with 100 replicates per scenario. (A) Distributions of posterior inclusion probabilities for instrument-specific pleiotropic effects across simulations with 0, 5, 10, 15, or 20 pleiotropic instruments among 20 genetic instruments. (B) Comparison of pleiotropic effect estimation error between CaPri and MR Egger across simulations with different numbers of pleiotropic instruments. Estimation error was calculated as the absolute difference between the estimated and ground-truth pleiotropic effects. Boxplots show the median (horizontal line), interquartile range (box), and values within 1.5 times the interquartile range from the lower and upper quartiles (whiskers).

When pleiotropic instruments were introduced, the PIP distribution became strongly bimodal, with distinct peaks near 0 and 1 corresponding to non-pleiotropic and pleiotropic instruments, respectively (**Figure 3A**). Under the primary simulation setting, sensitivity was 1.00 across all proportions of pleiotropic instruments. With five pleiotropic instruments, specificity and precision were 0.98 and 0.95, respectively, using PIP > 0.95, and both reached 1.00 when 10 or more instruments were pleiotropic. Results were nearly identical using the more stringent PIP > 0.99 threshold. Across simulations varying causal effect size, pleiotropic effect size, and confounding strength, sensitivity remained 1.00, while specificity ranged from 0.97 to 1.00 and precision from 0.93 to 1.00, demonstrating robust identification of pleiotropic instruments across the evaluated settings (**Supplementary Table 1**).

CaPri also accurately estimated the magnitude of the shared pleiotropic effect across different proportions of pleiotropic instruments (**Figure 3B**), consistent with its explicit modeling of instrument-specific membership in the pleiotropic component. In contrast, MR Egger became progressively more accurate as the proportion of instruments sharing the pleiotropic effect increased and achieved accuracy comparable to CaPri only when all instruments were pleiotropic.

### Impact of outcome GWAS sample size and prior settings

We next evaluated the impact of outcome GWAS sample size on the performance of CaPri. CaPri maintained strong performance with an outcome GWAS sample size of *n*_*Y*_ =10000, with the inferred PP.causal exceeding 0.99 in all simulation replicates across all proportions of pleiotropic instruments and the causal effect estimation error remaining low (**Supplementary Figure 4; Supplementary Table 1**). Identification of pleiotropic instruments also remained highly accurate, with sensitivity, specificity, and precision approaching 1.00 across the evaluated settings.

In contrast, performance of CaPri declined substantially when the outcome GWAS sample size was further reduced to *n*_*Y*_ =2000. The proportion of simulation replicates with PP.causal > 0.95 decreased substantially, ranging from 0% to 41% across the evaluated pleiotropy settings (**Supplementary Table 1**). Correspondingly, causal effect estimation error increased substantially. Sensitivity for identifying pleiotropic instruments also decreased, particularly when pleiotropy was widespread, declining to 0.62 when all instruments had pleiotropic effects, although specificity and precision remained high.

The influence of prior specification depended strongly on the outcome GWAS sample size (**Supplementary Figures 5-7; Supplementary Table 1**). With outcome GWAS sample sizes of *n*_*Y*_ =50000 and *n*_*Y*_ =10000, posterior estimates were highly similar across a broad range of prior specifications. In contrast, prior specification had a greater influence at *n*_*Y*_ =2000, when the likelihood provided substantially weaker evidence for distinguishing the causal and pleiotropic components. This pattern was most evident for the prior probability of a causal effect (*π*_δ_). In simulations with a true causal effect, increasing *π*_δ_directly resulted in higher PP.causal and lower causal effect estimation error at *n*_*Y*_ =2000 (**Supplementary Figure 5; Supplementary Table 1**).

### Evaluation of CaPri in real data analyses

We evaluated CaPri using negative and positive control exposure-outcome pairs. In the negative control analyses (**Supplementary Table 2**), CaPri provided little evidence for a causal effect of any of the ten exposures on self-reported tanning ability, with PP.causal < 0.01 for all exposure-outcome pairs (**Figure 4A; Supplementary Table 3**). In contrast, IVW, median-based methods, and MR PRESSO yielded significant associations for several negative control pairs involving asthma, body mass index, waist-to-hip ratio, or years of schooling.

**Figure 4.**
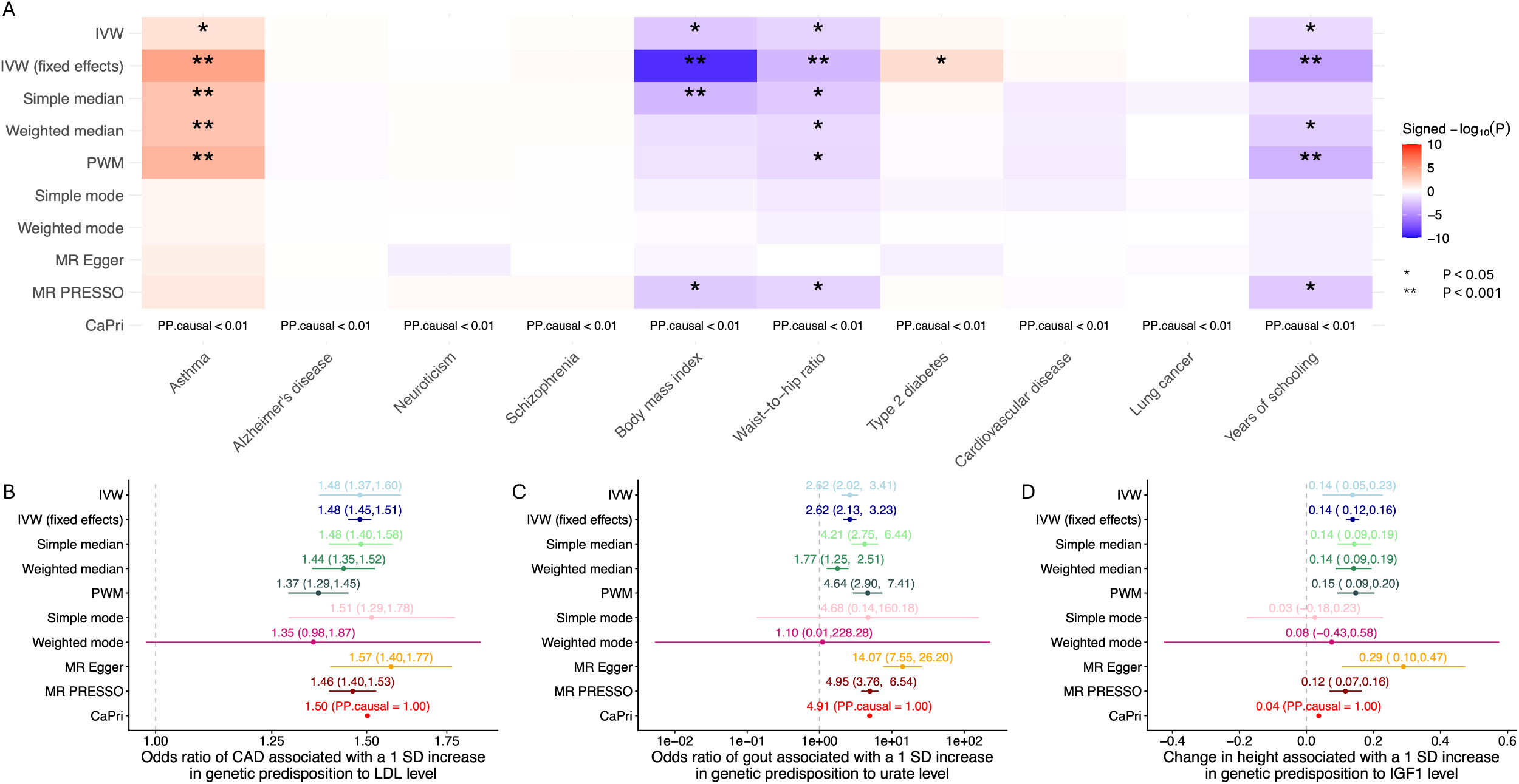
Negative and positive control analyses. (A) Heatmap of results from negative control analyses across 10 exposures and different MR methods. For conventional MR methods, colors represent statistical significance based on P-values, with red indicating positive effect estimates and blue indicating negative effect estimates. (B–D) Positive control analyses of (B) low-density lipoprotein (LDL) cholesterol on coronary artery disease (CAD), (C) urate on gout, and (D) insulin-like growth factor 1 (IGF-1) on height. Dots represent point estimates for conventional MR methods and the posterior mean causal effect for CaPri. Error bars represent 95% confidence intervals for conventional MR methods.

We next evaluated three positive control pairs with established causal or biological relationships, encompassing LDL cholesterol with CAD, serum urate with gout, and IGF-1 with height (**Supplementary Table 4**). CaPri provided strong evidence for a causal effect for all three pairs, with PP.causal approaching 1.00 (**Figure 4B-D; Supplementary Table 5**). These findings were consistent with IVW, median-based methods, MR Egger, and MR PRESSO, which also identified significant associations for all three positive controls. In contrast, mode-based methods had wider confidence intervals and did not consistently identify these established relationships, although their point estimates were in the expected direction.

Finally, we applied CaPri to investigate the relationship between alcohol consumption and CAD. All evaluated MR methods produced positive causal effect estimates, although the strength of statistical evidence differed substantially across methods (**Figure 5A; Supplementary Table 5**). CaPri estimated an OR of 1.38 for CAD per unit increase in genetically predicted log-transformed drinks per week, with PP.causal of 1.00. The corresponding estimates were OR = 1.16 (95% CI, 0.93-1.44; P = 0.19) for IVW, OR = 1.39 (95% CI, 1.22-1.59; P = 9.5×10^−7^) for weighted median, OR = 1.40 (95% CI, 1.22–1.60; P = 8.9×10^−7^) for penalized weighted median, OR = 1.36 (95% CI, 0.34– 5.51; P = 0.67) for weighted mode, OR = 1.47 (95% CI, 1.05–2.04; P = 0.03) for MR Egger, and OR = 1.15 (95% CI, 0.96–1.37; P = 0.14) for MR PRESSO.

CaPri additionally identified two instruments with high posterior probabilities of contributing to the pleiotropic component, which was negatively associated with CAD risk (**Figure 5B; Supplementary Table 5**). Notably, rs28929474 is a missense variant (NP_000286.3:p.Glu366Lys) in *SERPINA1*, which encodes alpha-1 antitrypsin, and represents the pathogenic Z allele associated with alpha-1 antitrypsin deficiency^52,53^. This variant has well-established associations with emphysema, chronic obstructive pulmonary disease, and liver disease^54,55^. One potential explanation for the negative pleiotropic effect of rs28929474 on CAD is a competing-risk mechanism, whereby carriers who develop severe pulmonary or hepatic disease may experience substantial morbidity or mortality before developing CAD, potentially resulting in an apparent protective association with CAD that does not operate through alcohol consumption. The identification of rs28929474 therefore provides a biologically plausible example of an instrument whose association with CAD may reflect a pathway distinct from the causal effect of alcohol consumption.

**Figure 5.**
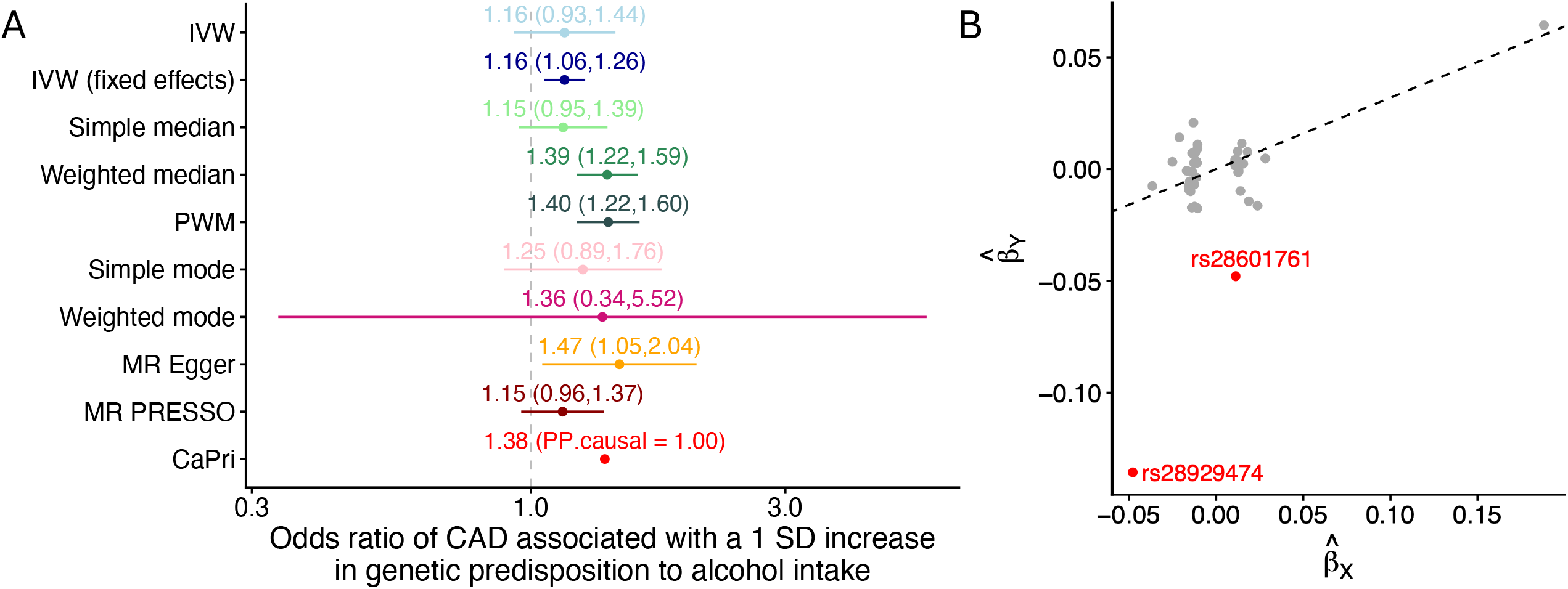
Mendelian randomization analysis of alcohol intake and coronary artery disease. (A) Estimated effect of alcohol intake on coronary artery disease (CAD) across MR methods. Dots represent point estimates for conventional MR methods and the posterior mean causal effect for CaPri. Error bars represent 95% confidence intervals for conventional MR methods. (B) Instrument-outcome associations 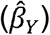 plotted against instrument-exposure associations 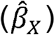. Each dot represents one genetic instrument, and the slope of the dashed line represents the posterior mean causal effect estimated by CaPri. Red dots indicate instruments identified as pleiotropic by CaPri, with their rsIDs indicated.

## Discussion

In this study, we developed CaPri, a Bayesian MR framework that jointly estimates causal and pleiotropic effects, providing the posterior probability and estimated magnitude of the causal effect, instrument-specific PIP for pleiotropy, and the estimated shared pleiotropic effect. CaPri retains the relevance and independence assumptions of MR while relaxing the exclusion restriction assumption by allowing a potentially arbitrary proportion of instruments to have direct effects on the outcome. To distinguish these effects from the causal component, CaPri assumes that pleiotropic instruments share a common directional effect.

Across simulations, CaPri maintained appropriate false positive control regardless of the proportion of pleiotropic instruments and accurately estimated causal effects when a true effect was present. Its performance was comparable to IVW in the absence of pleiotropy and to MR Egger when all instruments shared the pleiotropic effect, while remaining accurate when only a subset of instruments was pleiotropic. CaPri also accurately identified pleiotropic instruments and estimated the magnitude of the shared pleiotropic effect. These findings were consistent across different causal effect sizes, pleiotropic effect sizes, and strengths of exposure-outcome confounding. In contrast, median- and mode-based methods deteriorated when their assumptions regarding the proportion or clustering of valid instruments were violated^16–18^, whereas MR PRESSO was less well suited to settings in which pleiotropy represented a shared directional component rather than isolated outliers^19^. These results reinforce that the performance of pleiotropy-robust MR methods depends on how well their identifying assumptions reflect the underlying pleiotropic architecture.

Outcome GWAS sample size was an important determinant of CaPri performance. With outcome GWAS sample sizes of 50,000 or 10,000, CaPri maintained strong performance and showed little sensitivity to a broad range of prior specifications. In contrast, performance declined at a sample size of 2,000, with lower detection of true causal effects, greater estimation error, and reduced sensitivity for identifying pleiotropic instruments. Prior specification also became more influential, particularly the prior probability of a causal effect. This is expected for a Bayesian method, as the likelihood increasingly dominates the prior as statistical information increases. We therefore recommend prioritizing the use of large-scale outcome GWAS when applying CaPri, particularly when limited prior dependence is desired.

The real data analyses further supported the practical utility of CaPri. CaPri reported low PP.causal across all negative control analyses and strong posterior support for the three established positive control relationships. In the exploratory analysis of alcohol consumption and CAD, CaPri supported a positive causal effect while also identifying potentially meaningful pleiotropic instruments, including rs28929474 in *SERPINA1*. This example illustrates how CaPri may complement causal effect estimation by identifying instruments whose outcome associations may arise through distinct pleiotropic pathways.

Several limitations should be noted. First, CaPri currently models a single shared directional pleiotropic component. More heterogeneous pleiotropic architectures involving multiple distinct effects may not be adequately captured. Extending the framework to multiple pleiotropic components is possible but requires stronger regularization to ensure identifiability, which can substantially increase sensitivity to prior specification. Developing extensions that accommodate multiple pleiotropic components while limiting prior dependence remains an important direction for future work. Second, CaPri assumes that directional pleiotropy arises from either correlated or uncorrelated pleiotropy, but not both simultaneously. When correlated and uncorrelated pleiotropy coexist, their effects may not be adequately represented by the current model and would require additional identifying assumptions to distinguish^56^. Third, allele orientation is important because CaPri estimates a signed shared pleiotropic effect. In the real data analyses, instruments were oriented to the minor allele, which also reduces mechanical collinearity between the causal and pleiotropic components. However, the minor allele has no privileged biological direction, and heterogeneous pleiotropic effects with opposing directions may be less well captured. Finally, CaPri treats instrument-exposure associations as fixed and does not model their estimation uncertainty. Weak instruments may therefore introduce bias, although this concern may be mitigated when exposure associations are estimated in large-scale GWAS.

In conclusion, CaPri provides a complementary approach to MR inference in the presence of horizontal pleiotropy, allowing a potentially arbitrary proportion of genetic instruments to have pleiotropic effects. Our simulations and real data analyses demonstrate its ability to provide reliable causal inference and identify potentially pleiotropic instruments when its underlying assumptions are reasonably satisfied. By offering a distinct set of assumptions from existing pleiotropy-robust methods, CaPri can strengthen triangulation of causal evidence and improve the robustness of MR analyses.

## Acknowledgements

This research was supported in part by the Social Science Computing Center at the University of Wisconsin-Madison, which provided computing infrastructure and technical consulting services.

## Data and availability

CaPri is openly available at https://github.com/OuRen0104/CaPri.

