## Supplementary Figures for "CaPri: Bayesian Mendelian randomization for joint estimation of causal and pleiotropic effects"

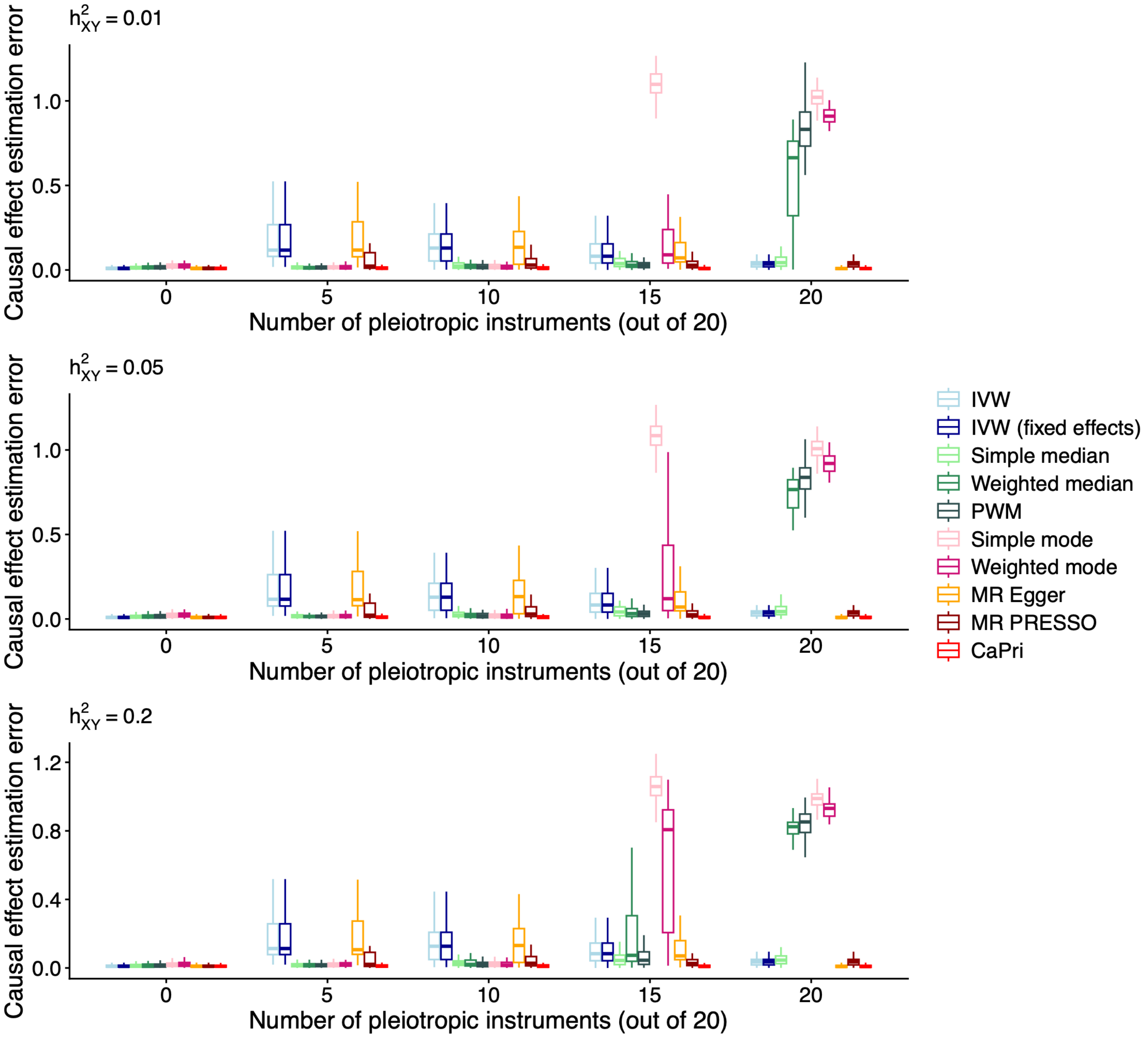


Supplementary Figure 1. Comparison of CaPri and other Mendelian randomization methods for causal effect inference across different causal effect sizes. Results were obtained from the primary simulation setting with 100 replicates per scenario. Results are shown across simulations with 0, 5, 10, 15, or 20 pleiotropic instruments among 20 genetic instruments. Boxplots show the median (horizontal line), interquartile range (box), and values within 1.5 times the interquartile range from the lower and upper quartiles (whiskers).

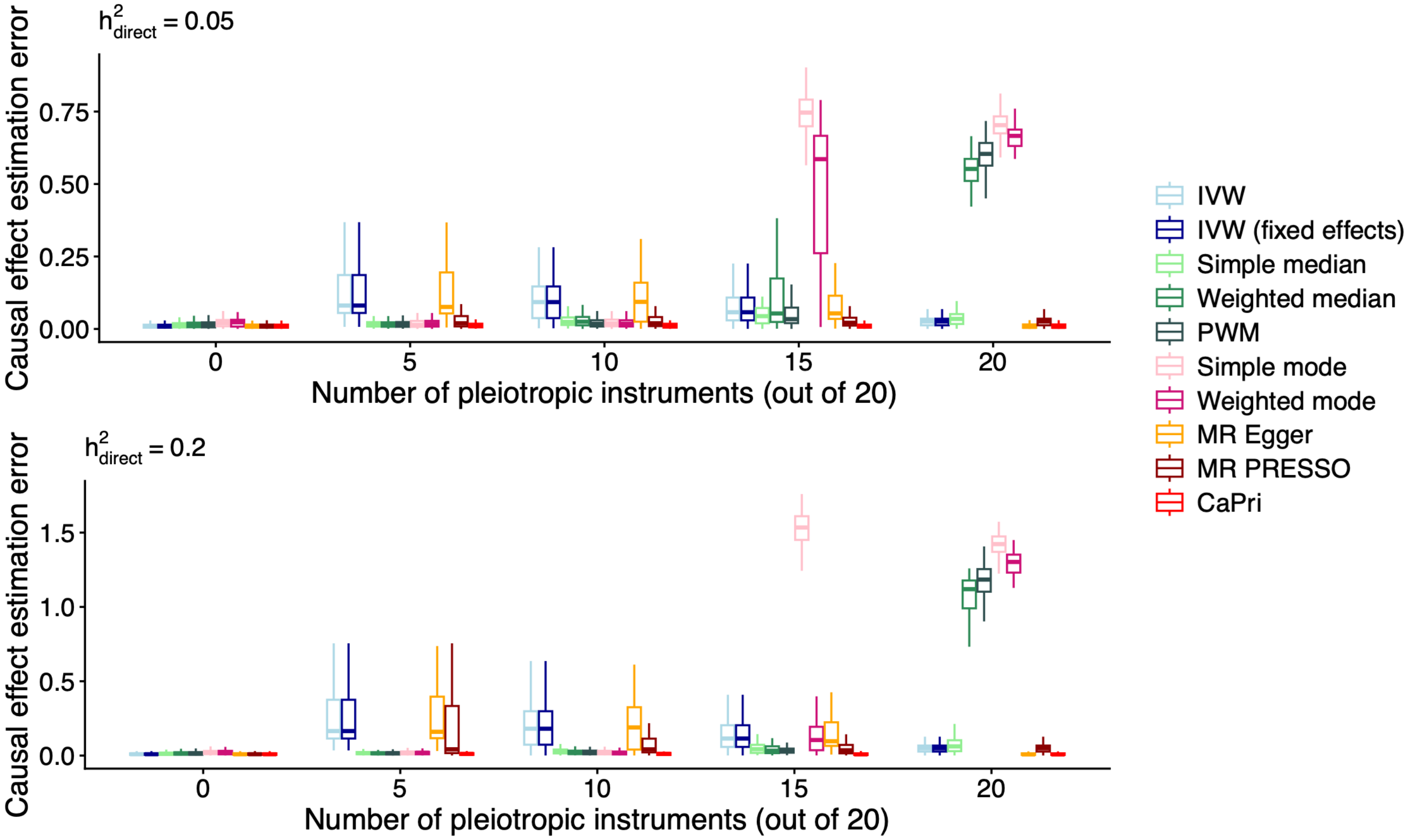


Supplementary Figure 2. Comparison of CaPri and other Mendelian randomization methods for causal effect inference across different pleiotropic effect sizes. Results were obtained from the primary simulation setting with 100 replicates per scenario. Results are shown across simulations with 0, 5, 10, 15, or 20 pleiotropic instruments among 20 genetic instruments. Boxplots show the median (horizontal line), interquartile range (box), and values within 1.5 times the interquartile range from the lower and upper quartiles (whiskers).

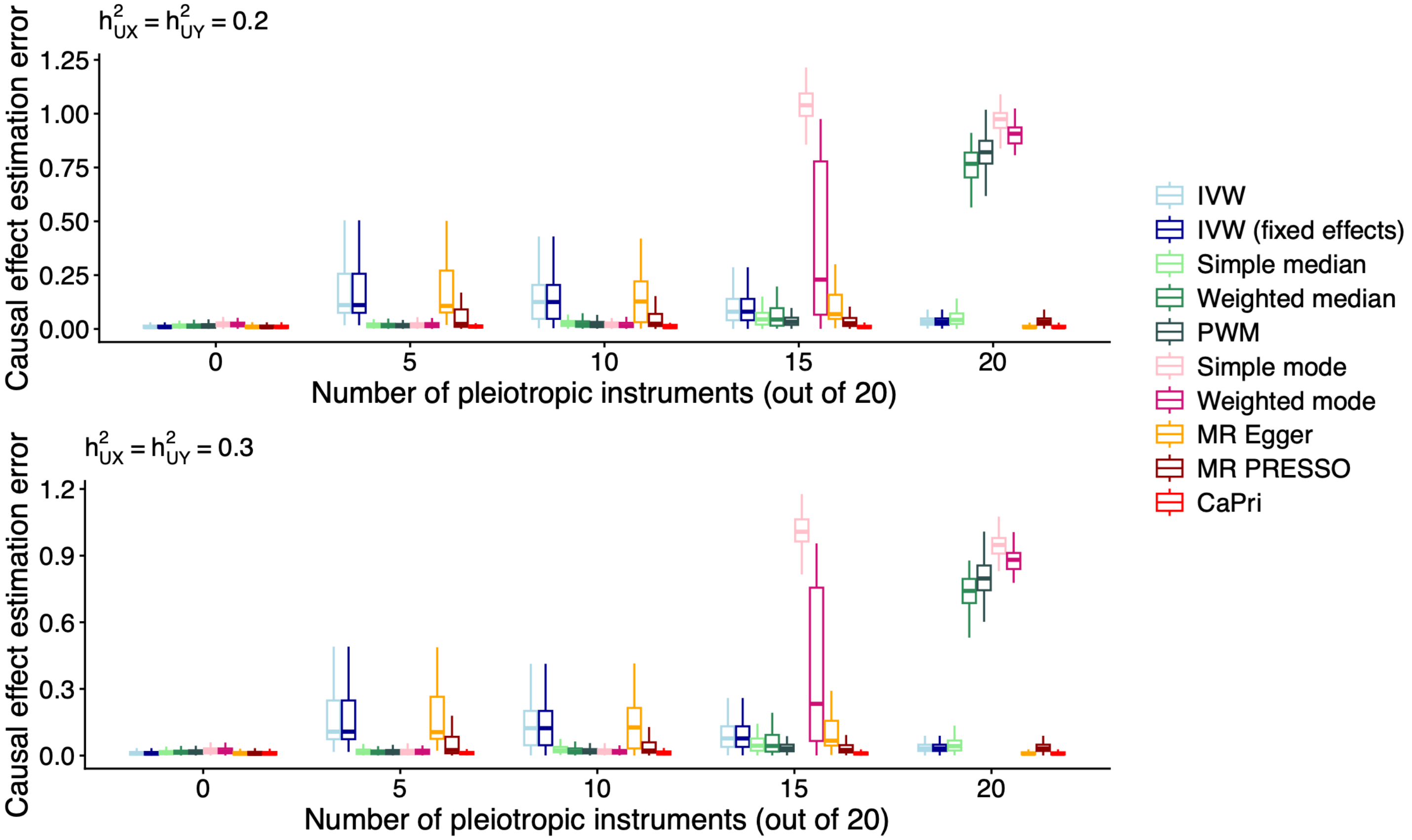


Supplementary Figure 3. Comparison of CaPri and other Mendelian randomization methods for causal effect inference across different magnitudes of exposure-outcome confounding. Results were obtained from the primary simulation setting with 100 replicates per scenario. Results are shown across simulations with 0, 5, 10, 15, or 20 pleiotropic instruments among 20 genetic instruments. Boxplots show the median (horizontal line), interquartile range (box), and values within 1.5 times the interquartile range from the lower and upper quartiles (whiskers).

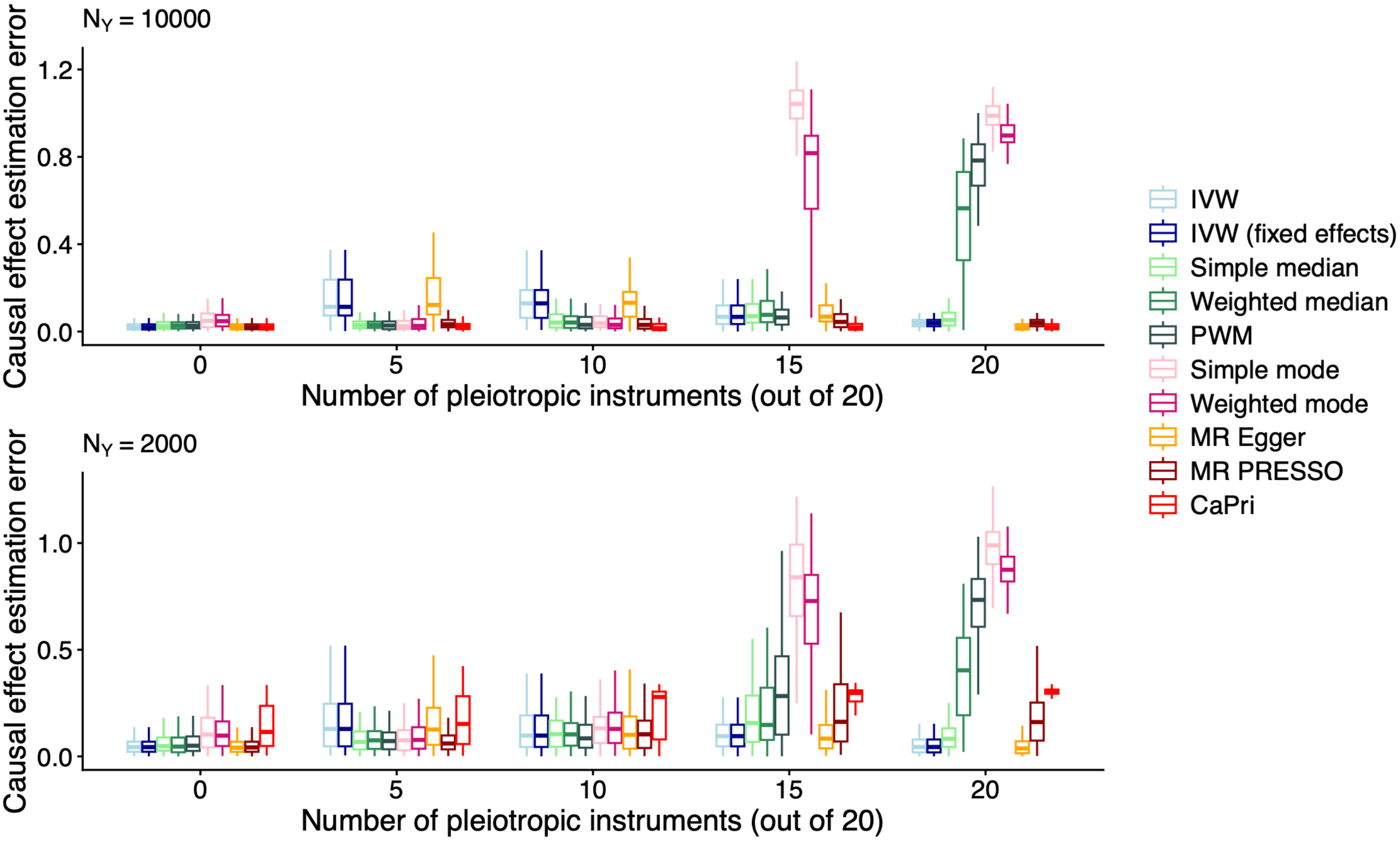


Supplementary Figure 4. Comparison of CaPri and other Mendelian randomization methods for causal effect inference across different outcome GWAS sample sizes. Results were obtained from the primary simulation setting with 100 replicates per scenario. Results are shown across simulations with 0, 5, 10, 15, or 20 pleiotropic instruments among 20 genetic instruments. Boxplots show the median (horizontal line), interquartile range (box), and values within 1.5 times the interquartile range from the lower and upper quartiles (whiskers).

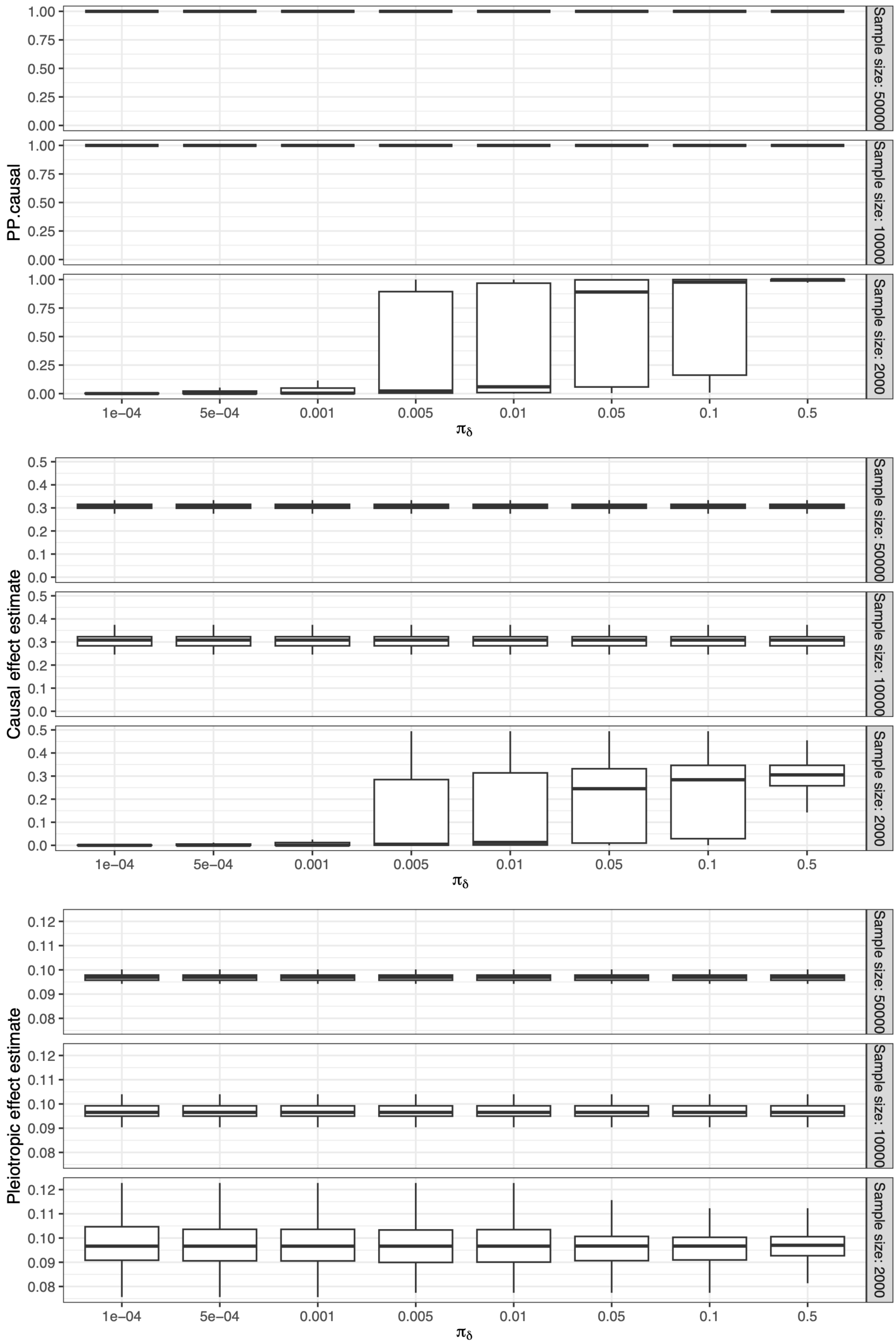


Supplementary Figure 5. Distribution of CaPri posterior estimates across different outcome GWAS sample sizes and prior probabilities of a causal effect. Results are shown for simulations in which 10 of 20 genetic instruments were pleiotropic. Boxplots show the distribution of each posterior estimate across 100 simulation replicates for each setting, with the median indicated by the horizontal line, the interquartile range by the box, and values within 1.5 times the interquartile range from the lower and upper quartiles by the whiskers.

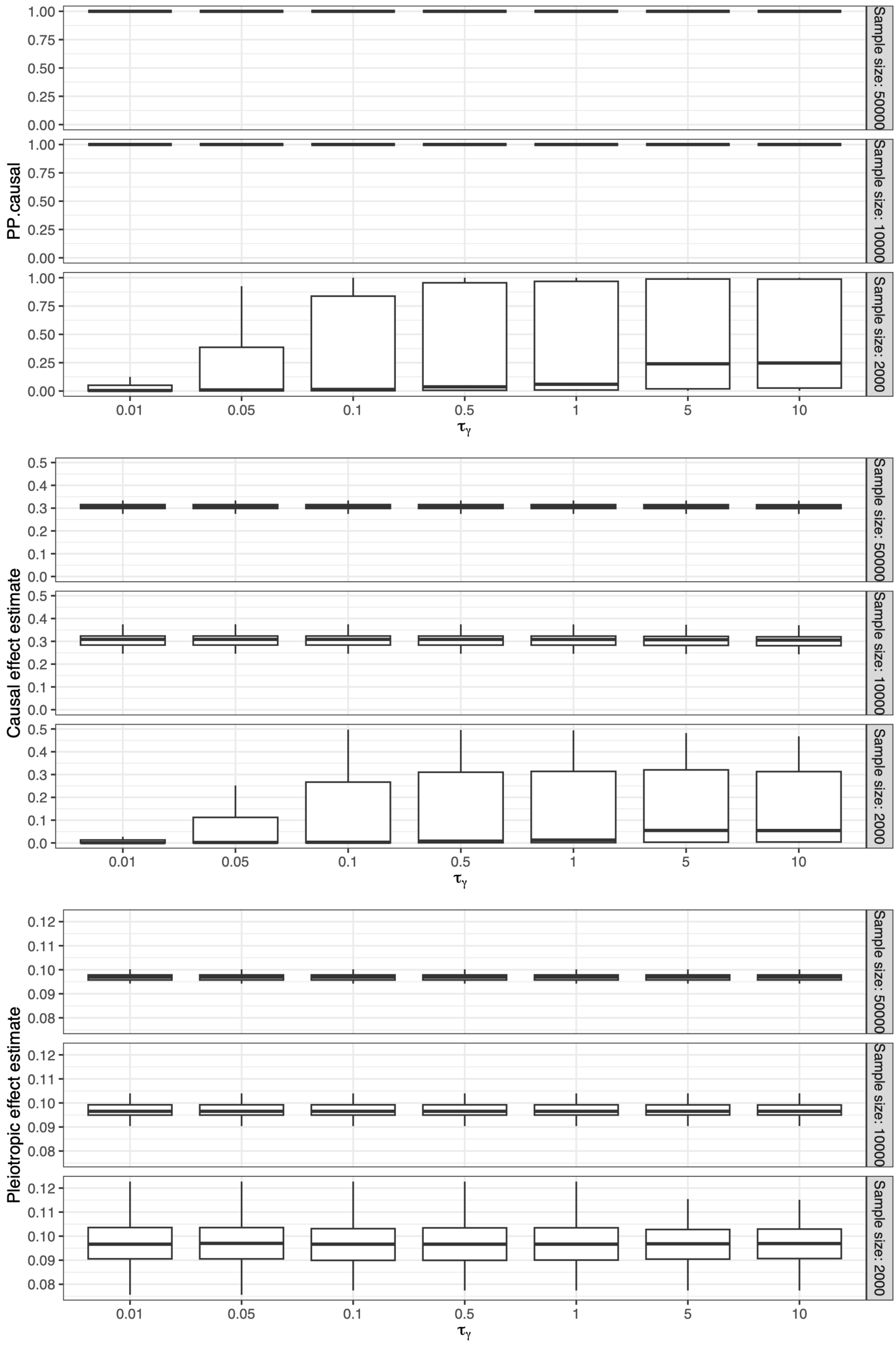


Supplementary Figure 6. Distribution of CaPri posterior estimates across different outcome GWAS sample sizes and prior precisions of the causal effect. Results are shown for simulations in which 10 of 20 genetic instruments were pleiotropic. Boxplots show the distribution of each posterior estimate across 100 simulation replicates for each setting, with the median indicated by the horizontal line, the interquartile range by the box, and values within 1.5 times the interquartile range from the lower and upper quartiles by the whiskers.

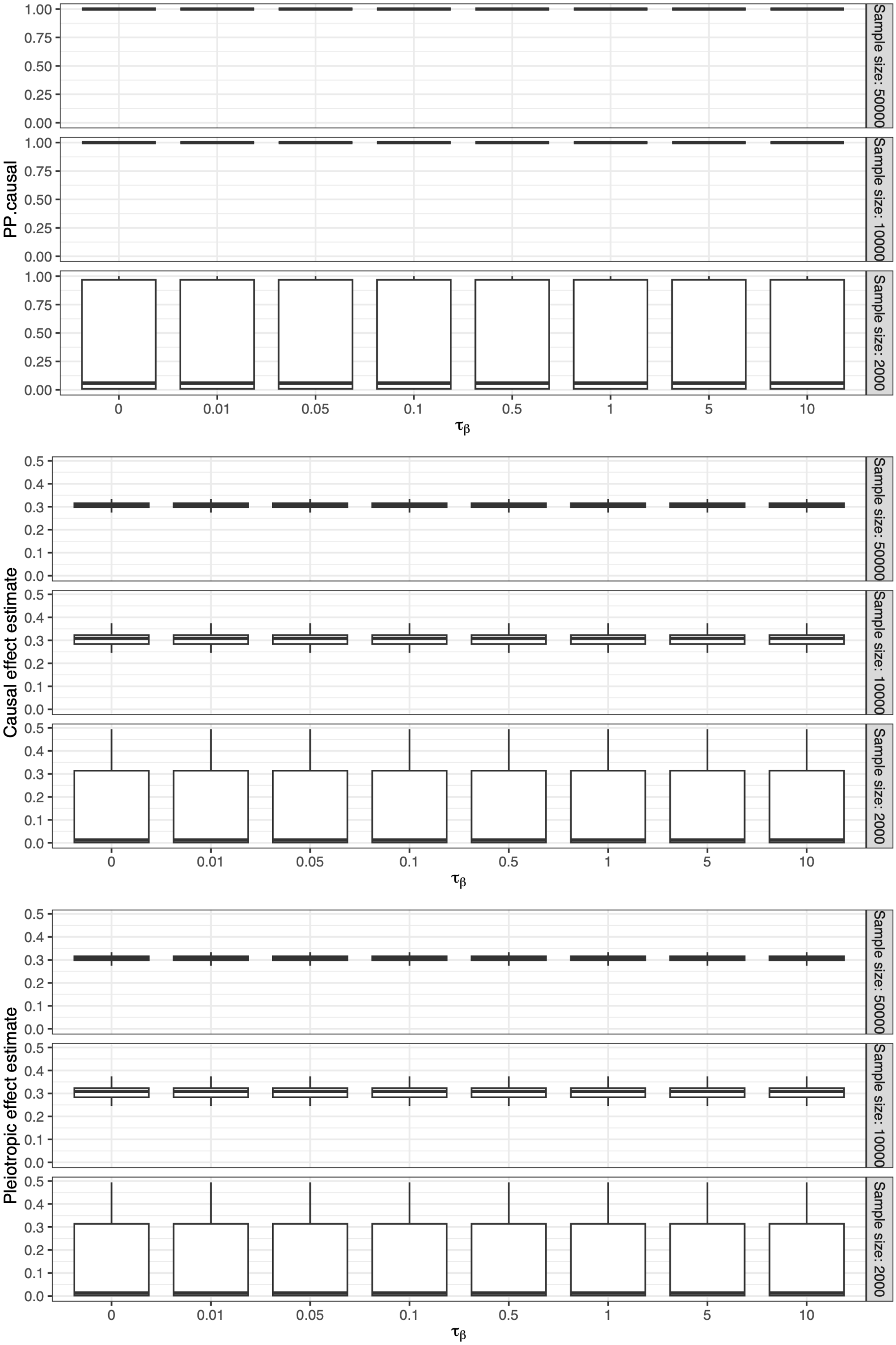


Supplementary Figure 7. Distribution of CaPri posterior estimates across different outcome GWAS sample sizes and prior precisions of the pleiotropic effect. Results are shown for simulations in which 10 of 20 genetic instruments were pleiotropic. Boxplots show the distribution of each posterior estimate across 100 simulation replicates for each setting, with the median indicated by the horizontal line, the interquartile range by the box, and values within 1.5 times the interquartile range from the lower and upper quartiles by the whiskers.
