## Supplementary Notes for "CaPri: Bayesian Mendelian randomization for joint estimation of causal and pleiotropic effects"

### Summary-statistics model

Consider $K$ independent genetic instruments. For the $k$-th instrument, let $\hat{\beta}_{X,k}$ and $\hat{\beta}_{Y,k}$ denote the estimated instrument-exposure and instrument-outcome associations, respectively. The summary-statistics implementation of CaPri treats $\hat{\beta}_{X,k}$ as fixed and models $\hat{\beta}_{Y,k}$ as

$$\hat{\beta}_{Y,k} = \gamma\hat{\beta}_{X,k} + c_{kY}\beta_Y + \epsilon_k, \qquad k = 1, \ldots, K,$$

where

$$\epsilon_k \sim \mathcal{N}\left(0, \frac{\sigma_Y^2}{n_Y}\right).$$

Here, $n_Y$ is the outcome GWAS sample size, $\gamma$ is the causal effect of the exposure on the outcome, $c_{kY}$ indicates whether instrument $k$ contributes to the pleiotropic component, and $\beta_Y$ is the shared directional pleiotropic effect. The corresponding likelihood is

$$p(\hat{\boldsymbol{\beta}}_Y \mid \hat{\boldsymbol{\beta}}_X, \gamma, \beta_Y, \boldsymbol{c}_Y) = \prod_{k=1}^{K} \mathcal{N}\left(\hat{\beta}_{Y,k}; \gamma\hat{\beta}_{X,k} + c_{kY}\beta_Y, \frac{\sigma_Y^2}{n_Y}\right).$$

Equivalently, up to a constant independent of the model parameters,

$$\log p(\hat{\boldsymbol{\beta}}_Y \mid \hat{\boldsymbol{\beta}}_X, \gamma, \beta_Y, \boldsymbol{c}_Y) = -\frac{n_Y}{2\sigma_Y^2}\sum_{k=1}^{K}\left(\hat{\beta}_{Y,k} - \gamma\hat{\beta}_{X,k} - c_{kY}\beta_Y\right)^2.$$

### Prior distributions

CaPri introduces a binary variable $\delta$ indicating whether a causal effect is present,

$$\delta \sim \text{Bernoulli}(\pi_\delta).$$

Conditional on $\delta$, the causal effect follows a spike-and-slab prior,

$$\gamma \mid \delta = \begin{cases} 0, & \delta = 0, \\ \mathcal{N}(0, \tau_\gamma^{-1}), & \delta = 1. \end{cases}$$

Thus, $\pi_\delta$ specifies the prior probability that the exposure has a causal effect on the outcome, and $\tau_\gamma$ is the prior precision of the causal effect under the slab component.
For each genetic instrument,

$$c_{kY} \sim \text{Bernoulli}(\pi_c),$$

where $\pi_c$ is the prior probability that an instrument contributes to the pleiotropic component.
The prior kernel for the shared pleiotropic effect is

$$p(\beta_Y) \propto \exp\left(-\frac{1}{2}\tau_\beta\beta_Y^2\right),$$

where $\tau_\beta$ is the prior precision. When $\tau_\beta > 0$, this corresponds to

$$\beta_Y \sim \mathcal{N}(0, \tau_\beta^{-1}).$$

The default setting $\tau_\beta = 0$ corresponds to the flat-prior limit used in the variational updates.

### Variational approximation

Exact posterior inference requires averaging over the $2^K$ possible configurations of the pleiotropy indicators in addition to the causal spike-and-slab component. CaPri therefore uses coordinate-ascent variational inference.

The variational distribution is factorized as

$$q(\beta_Y, \boldsymbol{c}_Y, \delta, \gamma) = q(\beta_Y) \left\{ \prod_{k=1}^{K} q(c_{kY}) \right\} q(\delta, \gamma),$$

where the causal indicator and causal effect are retained as a joint spike-and-slab factor. Specifically,

$$q(\beta_Y) = \mathcal{N}\left(\mu_\beta, (\tau_\beta^*)^{-1}\right),$$

$$q(c_{kY} = 1) = p_k,$$

$$q(\delta = 1) = q_\delta,$$

and

$$q(\gamma \mid \delta) = \begin{cases} 0, & \delta = 0, \\ \mathcal{N}\left(\mu_\gamma, (\tau_\gamma^*)^{-1}\right), & \delta = 1. \end{cases}$$

The quantities $p_k$ are the posterior inclusion probabilities for the instrument-specific pleiotropy indicators. The posterior moments required for the coordinate updates are

$$E_q[\gamma] = q_\delta \mu_\gamma,$$

$$E_q[\beta_Y] = \mu_\beta,$$

and

$$E_q[\beta_Y^2] = \mu_\beta^2 + \frac{1}{\tau_\beta^*}.$$

For convenience, define

$$\bar{\gamma} = E_q[\gamma] = q_\delta \mu_\gamma.$$

### Variational update for the shared pleiotropic effect

Holding the remaining variational factors fixed, the optimal variational distribution for $\beta_Y$ satisfies

$$\log q^*(\beta_Y) = E_{-\beta_Y}\left[\log p(\hat{\boldsymbol{\beta}}_Y, \beta_Y, \boldsymbol{c}_Y, \delta, \gamma \mid \hat{\boldsymbol{\beta}}_X)\right] + \text{constant}.$$

Define the residual with respect to the current posterior mean causal effect as

$$r_{\beta,k} = \hat{\beta}_{Y,k} - \bar{\gamma}\hat{\beta}_{X,k}.$$

Because $c_{kY}$ is binary,

$$E_q[c_{kY}] = E_q[c_{kY}^2] = p_k.$$

Retaining terms that depend on $\beta_Y$ gives

$$\log q^*(\beta_Y) = -\frac{1}{2}\left[\tau_\beta + \frac{n_Y}{\sigma_Y^2}\sum_{k=1}^{K} p_k\right]\beta_Y^2 + \frac{n_Y}{\sigma_Y^2}\left[\sum_{k=1}^{K} p_k r_{\beta,k}\right]\beta_Y + \text{constant}.$$

Therefore,

$$\tau_\beta^* = \tau_\beta + \frac{n_Y}{\sigma_Y^2}\sum_{k=1}^{K} p_k,$$

and

$$\mu_\beta = \frac{n_Y \sum_{k=1}^{K} p_k \left(\hat{\beta}_{Y,k} - \bar{\gamma}\hat{\beta}_{X,k}\right)}{\sigma_Y^2 \tau_\beta^*}.$$

The second posterior moment of the shared pleiotropic effect is subsequently calculated as

$$E_q[\beta_Y^2] = \mu_\beta^2 + \frac{1}{\tau_\beta^*}.$$

### Variational update for instrument-specific pleiotropy

For each instrument $k$, the variational posterior for $c_{kY}$ is Bernoulli. Its posterior log-odds can be written as the prior log-odds plus the expected difference in log likelihood between $c_{kY} = 1$ and $c_{kY} = 0$,

$$\log \frac{p_k}{1-p_k} = \log \frac{\pi_c}{1-\pi_c} + E_{-c_{kY}}\left[\log \frac{p(\hat{\beta}_{Y,k} \mid c_{kY} = 1)}{p(\hat{\beta}_{Y,k} \mid c_{kY} = 0)}\right].$$

Define

$$r_{c,k} = \hat{\beta}_{Y,k} - \bar{\gamma}\hat{\beta}_{X,k}.$$

For $c_{kY} = 0$, the contribution of the pleiotropic component is zero. For $c_{kY} = 1$, the residual becomes $r_{c,k} - \beta_Y$. The difference in the expected squared residuals is therefore determined by

$$E_q\left[(r_{c,k} - \beta_Y)^2 - r_{c,k}^2\right] = -2\mu_\beta r_{c,k} + E_q[\beta_Y^2].$$

It follows that

$$\text{logit}(p_k) = \text{logit}(\pi_c) + \frac{n_Y}{\sigma_Y^2}\left[\mu_\beta\left(\hat{\beta}_{Y,k} - \bar{\gamma}\hat{\beta}_{X,k}\right) - \frac{1}{2}\left(\mu_\beta^2 + \frac{1}{\tau_\beta^*}\right)\right].$$

Thus,

$$p_k = \frac{1}{1+\exp(-a_k)},$$

where

$$a_k = \log \frac{\pi_c}{1-\pi_c} + \frac{n_Y}{\sigma_Y^2}\left[\mu_\beta\left(\hat{\beta}_{Y,k} - \bar{\gamma}\hat{\beta}_{X,k}\right) - \frac{1}{2}\left(\mu_\beta^2 + \frac{1}{\tau_\beta^*}\right)\right].$$

The resulting $p_k$ is reported as the posterior inclusion probability for pleiotropy for instrument $k$.

### Variational update for the causal effect

Conditional on $\delta = 1$, the optimal variational distribution for the causal effect is Gaussian. Define the expected pleiotropy-adjusted instrument-outcome association as

$$r_{\gamma,k} = \hat{\beta}_{Y,k} - p_k \mu_\beta.$$

The expected log joint distribution as a function of $\gamma$, conditional on $\delta = 1$, is

$$\log q^*(\gamma \mid \delta = 1) = -\frac{1}{2}\left[\tau_\gamma + \frac{n_Y}{\sigma_Y^2}\sum_{k=1}^{K}\hat{\beta}_{X,k}^2\right]\gamma^2 + \frac{n_Y}{\sigma_Y^2}\left[\sum_{k=1}^{K}\hat{\beta}_{X,k} r_{\gamma,k}\right]\gamma + \text{constant}.$$

Therefore,

$$\tau_\gamma^* = \tau_\gamma + \frac{n_Y}{\sigma_Y^2}\sum_{k=1}^{K}\hat{\beta}_{X,k}^2,$$

and

$$\mu_\gamma = \frac{n_Y \sum_{k=1}^{K}\hat{\beta}_{X,k}\left(\hat{\beta}_{Y,k} - p_k \mu_\beta\right)}{\sigma_Y^2 \tau_\gamma^*}.$$

Here, $\mu_\gamma$ represents the posterior mean causal effect conditional on inclusion of the causal component, that is, conditional on $\delta = 1$.

### Variational update for the causal indicator

The posterior probability that the causal component is present is obtained by comparing the spike at $\gamma = 0$ with the Gaussian slab. Conditional on the current variational estimates of the pleiotropic component, define

$$A_\gamma = \frac{n_Y}{\sigma_Y^2}\sum_{k=1}^{K}\hat{\beta}_{X,k}^2$$

and

$$B_\gamma = \frac{n_Y}{\sigma_Y^2}\sum_{k=1}^{K}\hat{\beta}_{X,k}\left(\hat{\beta}_{Y,k} - p_k \mu_\beta\right).$$

Then

$$\tau_\gamma^* = \tau_\gamma + A_\gamma$$

and

$$\mu_\gamma = \frac{B_\gamma}{\tau_\gamma^*}.$$

Completing the square in the Gaussian slab gives the coordinate-wise log evidence for inclusion of the causal component relative to the spike,

$$\frac{1}{2}\left[\log\left(\frac{\tau_\gamma}{\tau_\gamma^*}\right) + \tau_\gamma^* \mu_\gamma^2\right].$$

The posterior log-odds of a causal effect are therefore

$$\operatorname{logit}(q_\delta) = \operatorname{logit}(\pi_\delta) + \frac{1}{2}\left[\log\left(\frac{\tau_\gamma}{\tau_\gamma^*}\right) + \tau_\gamma^* \mu_\gamma^2\right].$$

Equivalently,

$$q_\delta = \frac{1}{1 + \exp(-a_\delta)},$$

where

$$a_\delta = \log \frac{\pi_\delta}{1-\pi_\delta} + \frac{1}{2}\left[\log\left(\frac{\tau_\gamma}{\tau_\gamma^*}\right) + \tau_\gamma^* \mu_\gamma^2\right].$$

The quantity

$$q_\delta = P_q(\delta = 1)$$

is reported as PP.causal.

### Posterior causal and pleiotropic effect estimates

Because the causal effect is exactly zero under the spike component,

$$E_q[\gamma] = P_q(\delta = 1) E_q[\gamma \mid \delta = 1] = q_\delta \mu_\gamma.$$

CaPri therefore reports

$$\hat{\gamma} = q_\delta \mu_\gamma$$

as the posterior mean causal effect.
The posterior mean shared pleiotropic effect is

$$\hat{\beta}_Y = E_q[\beta_Y] = \mu_\beta,$$

and the posterior probability that instrument $k$ contributes to the pleiotropic component is

$$\text{PIP}_k = P_q(c_{kY} = 1) = p_k.$$

Thus, the principal posterior quantities returned by CaPri are the posterior probability of a causal effect $q_\delta$, the posterior mean causal effect $q_\delta \mu_\gamma$, the posterior mean shared pleiotropic effect $\mu_\beta$, and the instrument-specific pleiotropy posterior inclusion probabilities $p_k$.

### Coordinate-ascent algorithm

The variational parameters are updated iteratively. The implementation initializes

$$q_\delta^{(0)} = 0.5, \qquad \mu_\gamma^{(0)} = 0, \qquad p_k^{(0)} = \pi_c, \qquad \mu_\beta^{(0)} = 0.$$

At each iteration, the following updates are performed in sequence.

1. Calculate the current posterior mean causal effect,

$$\bar{\gamma} = q_\delta \mu_\gamma.$$

2. Update the variational distribution of the shared pleiotropic effect,

$$\tau_\beta^* = \tau_\beta + \frac{n_Y}{\sigma_Y^2} \sum_{k=1}^{K} p_k,$$

$$\mu_\beta = \frac{n_Y \sum_{k=1}^{K} p_k \left(\hat{\beta}_{Y,k} - \bar{\gamma}\hat{\beta}_{X,k}\right)}{\sigma_Y^2 \tau_\beta^*}.$$

3. Calculate

$$E_q[\beta_Y^2] = \mu_\beta^2 + \frac{1}{\tau_\beta^*},$$

and update each pleiotropy posterior inclusion probability,

$$\text{logit}(p_k) = \text{logit}(\pi_c) + \frac{n_Y}{\sigma_Y^2}\left[\mu_\beta\left(\hat{\beta}_{Y,k} - \bar{\gamma}\hat{\beta}_{X,k}\right) - \frac{1}{2}E_q[\beta_Y^2]\right].$$

4. Update the variational distribution of the causal effect conditional on $\delta = 1$,

$$\tau_\gamma^* = \tau_\gamma + \frac{n_Y}{\sigma_Y^2}\sum_{k=1}^{K}\hat{\beta}_{X,k}^2,$$

$$\mu_\gamma = \frac{n_Y \sum_{k=1}^{K}\hat{\beta}_{X,k}\left(\hat{\beta}_{Y,k} - p_k\mu_\beta\right)}{\sigma_Y^2\tau_\gamma^*}.$$

5. Update the posterior probability of the causal component,

$$\text{logit}(q_\delta) = \text{logit}(\pi_\delta) + \frac{1}{2}\left[\log\left(\frac{\tau_\gamma}{\tau_\gamma^*}\right) + \tau_\gamma^*\mu_\gamma^2\right].$$

These steps are repeated until convergence. Because each update requires only sums over the $K$ instruments, the computational complexity of each iteration is linear in the number of instruments.

### Convergence criterion

Let $\theta^{(t)}$ denote the variational parameters at iteration $t$. The implementation monitors the maximum absolute change across the principal variational parameters,

$$d_\theta = \max\left\{\left|q_\delta^{(t)} - q_\delta^{(t-1)}\right|, \left|\mu_\gamma^{(t)} - \mu_\gamma^{(t-1)}\right|, \left|\mu_\beta^{(t)} - \mu_\beta^{(t-1)}\right|, \max_k\left|p_k^{(t)} - p_k^{(t-1)}\right|, \right.$$
$$\left.\left|\tau_\beta^{*(t)} - \tau_\beta^{*(t-1)}\right|, \left|\tau_\gamma^{*(t)} - \tau_\gamma^{*(t-1)}\right|, \left|q_\delta^{(t)}\mu_\gamma^{(t)} - q_\delta^{(t-1)}\mu_\gamma^{(t-1)}\right|\right\}.$$

Convergence is declared when

$$d_\theta < 10^{-6}$$

for five consecutive iterations. The default maximum number of iterations is 2,000.

### Allele orientation

Because the pleiotropic component is directional, allele orientation determines the sign of the shared pleiotropic effect. CaPri therefore allows summary statistics to be oriented to the minor allele. When minor-allele orientation is used, both the instrument-exposure and instrument-outcome associations are multiplied by $-1$ if the reported effect allele frequency for instrument $k$ exceeds 0.5. This preserves the relationship between the two associations while expressing all effects according to a common minor-allele convention. Importantly, minor-allele orientation avoids mechanically forcing all instrument-exposure associations to have the same sign, which could increase collinearity between the causal and shared directional pleiotropic components.
